# OpenSAFELY: Effectiveness of COVID-19 vaccination in children and adolescents

**DOI:** 10.1101/2024.05.20.24306810

**Authors:** Colm D Andrews, Edward P K Parker, Elsie Horne, Venexia Walker, Tom Palmer, Andrea L Schaffer, Amelia CA Green, Helen J Curtis, Alex J Walker, Lucy Bridges, Christopher Wood, Victoria Speed, Christopher Bates, Jonathan Cockburn, John Parry, Amir Mehrkar, Brian MacKenna, Sebastian CJ Bacon, Ben Goldacre, Miguel A Hernan, Jonathan AC Sterne, The OpenSAFELY Collaborative, William J Hulme

## Abstract

**Background:** Children and adolescents in England were offered BNT162b2 as part of the national COVID-19 vaccine roll out from September 2021. We assessed the safety and effectiveness of first and second dose BNT162b2 COVID-19 vaccination in children and adolescents in England.

**Methods:** With the approval of NHS England, we conducted an observational study in the OpenSAFELY-TPP database, including a) adolescents aged 12-15 years, and b) children aged 5-11 years and comparing individuals receiving i) first vaccination with unvaccinated controls and ii) second vaccination to single-vaccinated controls. We matched vaccinated individuals with controls on age, sex, region, and other important characteristics. Outcomes were positive SARS-CoV-2 test (adolescents only); COVID-19 A&E attendance; COVID-19 hospitalisation; COVID-19 critical care admission; COVID-19 death, with non-COVID-19 death and fractures as negative control outcomes and A&E attendance, unplanned hospitalisation, pericarditis, and myocarditis as safety outcomes.

**Results:** Amongst 820,926 previously unvaccinated adolescents, the incidence rate ratio (IRR) for positive SARS-CoV-2 test comparing vaccination with no vaccination was 0.74 (95% CI 0.72-0.75), although the 20-week risks were similar. The IRRs were 0.60 (0.37-0.97) for COVID-19 A&E attendance, 0.58 (0.38-0.89) for COVID-19 hospitalisation, 0.99 (0.93-1.06) for fractures, 0.89 (0.87-0.91) for A&E attendances and 0.88 (0.81-0.95) for unplanned hospitalisation. Amongst 441,858 adolescents who had received first vaccination IRRs comparing second dose with first dose only were 0.67 (0.65-0.69) for positive SARS-CoV-2 test, 1.00 (0.20-4.96) for COVID-19 A&E attendance, 0.60 (0.26-1.37) for COVID-19 hospitalisation, 0.94 (0.84-1.05) for fractures, 0.93 (0.89-0.98) for A&E attendance and 0.99 (0.86-1.13) for unplanned hospitalisation. Amongst 283,422 previously unvaccinated children and 132,462 children who had received a first vaccine dose, COVID-19-related outcomes were too rare to allow IRRs to be estimated precisely. A&E attendance and unplanned hospitalisation were slightly higher after first vaccination (IRRs versus no vaccination 1.05 (1.01-1.10) and 1.10 (0.95-1.26) respectively) but slightly lower after second vaccination (IRRs versus first dose 0.95 (0.86-1.05) and 0.78 (0.56-1.08) respectively). There were no COVID-19-related deaths in any group. Fewer than seven (exact number redacted) COVID-19-related critical care admissions occurred in the adolescent first dose vs unvaccinated cohort. Among both adolescents and children, myocarditis and pericarditis were documented only in the vaccinated groups, with rates of 27 and 10 cases/million after first and second doses respectively.

**Conclusion:** BNT162b2 vaccination in adolescents reduced COVID-19 A&E attendance and hospitalisation, although these outcomes were rare. Protection against positive SARS-CoV-2 tests was transient.

## Background

The UK’s COVID-19 vaccination programme was extended to adolescents aged 12-15 years on 20 September 2021, with an adult 30 µg dose of Pfizer-BioNTech (BNT162b2) vaccine licensed for use(1). Children aged 5-11 years were eligible from 4 April 2022, using a 10µg dose(2). Individuals considered high-risk, for example those with immunosuppressive conditions or living with a vulnerable adult, were eligible earlier (adolescents in August 2021(3) and children in January 2022(4)).

Authorisation in children and adolescents was based on phase II/III randomized trials (RCTs) showing high immunogenicity and efficacy against infection. However, protection against severe disease and safety endpoints were not assessed in RCTs(5). Multiple countries have reported rare cases of myocarditis and pericarditis following mRNA COVID-19 vaccines(6–8): these events are listed in BNT162b2 product information(9).

We used the OpenSAFELY-TPP database, covering 40% of English primary care practices and linked to national coronavirus surveillance, hospital episodes, and death registry data, to emulate a hypothetical target trial to evaluate the effectiveness of childhood COVID-19 vaccination against COVID-19 related and other outcomes.

## Methods

### Data source

All data were linked, stored and analysed securely using the OpenSAFELY platform, https://www.opensafely.org/, as part of the NHS England OpenSAFELY COVID-19 service. Data include pseudonymised data such as coded diagnoses, medications and physiological parameters. No free text data was included. All code is shared openly for review and re-use under MIT open license [https://github.com/opensafely/vaccine-effectiveness-in-kids]. Detailed pseudonymised patient data is potentially re-identifiable and therefore not shared. Primary care records managed by the GP software provider, TPP were linked to ONS death data and the Index of Multiple Deprivation through OpenSAFELY.

### Eligibility criteria, vaccination groups, and matching

We included a) all adolescents aged 12-15 years and b) all children aged 5-11 years on 31 August 2021, when age-based vaccine eligibility criteria were defined, who were not clinically vulnerable, as defined by the Joint Committee on Vaccination and Immunisation (JCVI) (clinically vulnerable individuals were eligible for vaccination prior to the start of the study entry period); had been continuously registered at a GP practice using TPP’s SystmOne clinical information system for 42 days; had no evidence of SARS-CoV-2 infection or COVID-19 disease within the 30 days before vaccination; and had complete information on sex, deprivation, ethnicity, and NHS region.

We estimated the effectiveness and safety of: i) first vaccine dose vs. no vaccination, and ii) a second dose vs. a single dose only by emulating a sequence of target trials. For the first vaccine dose, on each day of the study period, each eligible individual vaccinated with their first dose was matched 1:1 without replacement with an eligible individual who had not yet been vaccinated. Unvaccinated individuals were matched to at most one vaccinated individual, but were eligible for inclusion in the vaccination group later on if they were subsequently vaccinated. Follow-up of the vaccinated group included time after second vaccination. We used the same approach to assess effectiveness and safety of a second vaccine dose, amongst individuals who had received a first vaccine dose.

Matching criteria were: age within year on 31 August 2021 (i.e., in the same school-year); sex (male/female); region; evidence of prior infection (yes/no); prior tests in the preceding 26 weeks (0, 1-2, 3+) and prior non-COVID childhood vaccination (MMR: yes/no); all other childhood vaccines(10) (yes/no) and, for the second dose comparison, first vaccination date within 7 days.

### Outcomes

Five effectiveness outcomes were considered: positive SARS-CoV-2 test; COVID-19 A&E attendance; COVID-19 hospitalisation; COVID-19 critical care admission; COVID-19 death. Freely-available community testing for COVID-19 ended on 31 March 2022, and as non-high-risk children became eligible for vaccination in April 2022 the positive SARS-CoV-2 test was not considered for children (age 5-11 years). non-COVID-19 death, fractures and effectiveness in the first week after vaccination were considered as negative control outcomes(11). We also considered A&E attendance, unplanned hospitalisation, pericarditis and myocarditis as safety endpoints (Supplementary Box 1). Outcomes are described in Supplementary Box 1.

### Follow-up

Each individual was followed from assignment to a comparison group (‘time zero’) until the earliest of: end of freely-available community testing (positive SARS-CoV-2 test only); outcome; death; practice de-registration; 20 weeks; vaccination of the unvaccinated control (first dose group only) or second vaccination of the control (second dose group only).

### Statistical Analysis

We estimated period-specific incidence rates in each treatment group (number of events divided by person-time at risk) and derived incidence rate ratios and 95% CIs. We also derived 20-week risk differences (RD), and corresponding 95% CIs from the sum of squares of Greenwood standard errors. We estimated effectiveness separately according to whether there was evidence of prior SARS-CoV-2 infection (Supplementary Tables 1-4).

### Software, code, and reproducibility

Data management and analyses were conducted in Python version 3.8.10 and R version 4.0.5. Code for data management and analysis, as well as codelists, are archived online https://github.com/opensafely/vaccine-effectiveness-in-kids.

### Disclosure control

Any reported figures based on counts below 8 were redacted. Cumulative incidence plots are not reported for outcomes with fewer than 30 outcomes. To reduce re-identification risk, counts are reported to the nearest n∗6−3 (3,9,15,21,…). All derived statistics are based on these rounded counts, including cumulative incidence curves which are based on rounded numbers-at-risk and event counts.

## Results

### Adolescents

Of 513,192 eligible adolescents who were registered at a TPP practice and received a BNT162b2 vaccination during the study period, 410,463 (80%) were matched with unvaccinated controls. (Supplementary Figure 1, panel A). 220,929 (81%) of 271,440 eligible adolescents who received a second BNT162b2 vaccination were matched with single vaccinated controls (Supplementary Figure 1, panel B).

As expected, the matching factors, other than age, were identically distributed in the vaccinated and control groups at the start of follow-up for all of the study populations (Table 1). Over 60% of adolescents in the first vaccination group received a second vaccination. Most second vaccinations occurred at least 12 weeks after first vaccination (Supplementary Figure 2).

**Table 1:** Characteristics N (%) of matched participants on the day of study entry. Counts are based on values rounded up to the nearest n*6-3, for disclosure control.

|  |  | Adolescents |  |  |  | Children |  |  |  |
| --- | --- | --- | --- | --- | --- | --- | --- | --- | --- |
|  |  | First dose vs Unvaccinated |  | Second Dose vs Single Dose Only |  | First dose vs Unvaccinated |  | Second dose vs Single Dose Only |  |
|  |  | First dose<br>(N=410,463) | Unvaccinated<br>(N=410,463) | Second dose<br>(N=220,929) | Single dose only<br>(N=220,929) | First dose<br>(N=141,711) | Unvaccinated<br>(N=141,711) | Second dose<br>(N=66,231) | Single dose only<br>(N=66,231) |
| Age | 5 | - | - | - | - | 3,537 (2.5) | 2,457 (1.7) | 237 (0.4) | 153 (0.2) |
|  | 6 | - | - | - | - | 13,887 (9.8) | 13,905 (9.8) | 5,859 (8.8) | 5,811 (8.8) |
|  | 7 | - | - | - | - | 15,837 (11.2) | 15,591 (11.0) | 6,909 (10.4) | 6,843 (10.3) |
|  | 8 | - | - | - | - | 17,865 (12.6) | 18,435 (13.0) | 8,181 (12.4) | 8,187 (12.4) |
|  | 9 | - | - | - | - | 22,497 (15.9) | 21,759 (15.4) | 10,329 (15.6) | 10,293 (15.5) |
|  | 10 | - | - | - | - | 26,505 (18.7) | 26,187 (18.5) | 12,639 (19.1) | 12,633 (19.1) |
|  | 11 | - | - | - | - | 37,311 (26.3) | 31,473 (22.2) | 15,855 (23.9) | 15,783 (23.8) |
|  | 12 | 79,083 (19.3) | 78,669 (19.2) | 28,353 (12.8) | 25,401 (11.5) | 4,275 (3.0) | 11,901 (8.4) | 6,219 (9.4) | 6,531 (9.9) |
|  | 13 | 103,227 (25.1) | 104,391 (25.4) | 54,291 (24.6) | 55,527 (25.1) | - | - | 3 (0.0) | 0 (0.0) |
|  | 14 | 103,773 (25.3) | 103,527 (25.2) | 56,595 (25.6) | 56,829 (25.7) | - | - | - | - |
|  | 15 | 104,895 (25.6) | 105,753 (25.8) | 56,685 (25.7) | 59,901 (27.1) | - | - | - | - |
|  | 16 | 19,485 (4.7) | 18,123 (4.4) | 24,999 (11.3) | 23,265 (10.5) | - | - | - | - |
| Sex | Female | 205,173 (50.0) | 205,173 (50.0) | 110,121 (49.8) | 110,121 (49.8) | 70,611 (49.8) | 70,611 (49.8) | 33,159 (50.1) | 33,159 (50.1) |
|  | Male | 205,287 (50.0) | 205,287 (50.0) | 110,805 (50.2) | 110,805 (50.2) | 71,103 (50.2) | 71,103 (50.2) | 33,075 (49.9) | 33,075 (49.9) |
| Deprivation | 1 (most deprived) | 70,947 (17.3) | 70,947 (17.3) | 31,419 (14.2) | 31,419 (14.2) | 20,103 (14.2) | 20,103 (14.2) | 8,421 (12.7) | 8,421 (12.7) |
|  | 2 | 72,831 (17.7) | 72,831 (17.7) | 36,057 (16.3) | 36,057 (16.3) | 23,577 (16.6) | 23,577 (16.6) | 10,395 (15.7) | 10,395 (15.7) |
|  | 3 | 84,081 (20.5) | 84,081 (20.5) | 45,417 (20.6) | 45,417 (20.6) | 29,253 (20.6) | 29,253 (20.6) | 13,473 (20.3) | 13,473 (20.3) |
|  | 4 | 87,891 (21.4) | 87,891 (21.4) | 50,307 (22.8) | 50,307 (22.8) | 32,103 (22.7) | 32,103 (22.7) | 15,435 (23.3) | 15,435 (23.3) |
|  | 5 (least deprived) | 94,707 (23.1) | 94,707 (23.1) | 57,723 (26.1) | 57,723 (26.1) | 36,675 (25.9) | 36,675 (25.9) | 18,513 (28.0) | 18,513 (28.0) |
| Region | East of England | 99,471 (24.2) | 99,471 (24.2) | 55,581 (25.2) | 55,581 (25.2) | 35,019 (24.7) | 35,019 (24.7) | 17,493 (26.4) | 17,493 (26.4) |
|  | London | 18,519 (4.5) | 18,519 (4.5) | 8,697 (3.9) | 8,697 (3.9) | 6,147 (4.3) | 6,147 (4.3) | 2,271 (3.4) | 2,271 (3.4) |
|  | Midlands | 90,093 (21.9) | 90,093 (21.9) | 50,601 (22.9) | 50,601 (22.9) | 31,497 (22.2) | 31,497 (22.2) | 14,931 (22.5) | 14,931 (22.5) |
|  | North East and Yorkshire | 78,471 (19.1) | 78,471 (19.1) | 41,061 (18.6) | 41,061 (18.6) | 24,105 (17.0) | 24,105 (17.0) | 11,853 (17.9) | 11,853 (17.9) |
|  | North West | 36,057 (8.8) | 36,057 (8.8) | 18,333 (8.3) | 18,333 (8.3) | 10,005 (7.1) | 10,005 (7.1) | 3,981 (6.0) | 3,981 (6.0) |
|  | South East | 26,511 (6.5) | 26,511 (6.5) | 14,571 (6.6) | 14,571 (6.6) | 9,651 (6.8) | 9,651 (6.8) | 4,059 (6.1) | 4,059 (6.1) |
|  | South West | 61,335 (14.9) | 61,335 (14.9) | 32,079 (14.5) | 32,079 (14.5) | 25,275 (17.8) | 25,275 (17.8) | 11,637 (17.6) | 11,637 (17.6) |
| Number of SARS-CoV-2 tests | 0 | 85,515 (20.8) | 85,515 (20.8) | 39,795 (18.0) | 39,795 (18.0) | 52,503 (37.0) | 52,503 (37.0) | 48,969 (73.9) | 48,969 (73.9) |
|  | 1-2 | 108,261 (26.4) | 108,261 (26.4) | 62,487 (28.3) | 62,487 (28.3) | 51,897 (36.6) | 51,897 (36.6) | 9,963 (15.0) | 9,963 (15.0) |
|  | 3+ | 216,681 (52.8) | 216,681 (52.8) | 118,647 (53.7) | 118,647 (53.7) | 37,311 (26.3) | 37,311 (26.3) | 7,299 (11.0) | 7,299 (11.0) |
| Prior documented SARS-CoV-2 infection |  | 82,137 (20.0) | 82,137 (20.0) | 50,007 (22.6) | 50,007 (22.6) | 65,307 (46.1) | 65,307 (46.1) | 30,777 (46.5) | 30,777 (46.5) |

### Effectiveness of first dose in adolescents

In 95,641 person-years of potential follow-up, there were 56,496 positive SARS-CoV-2 tests; 72 COVID-19 A&E attendances; 90 COVID-19 hospitalisations, of which 3 included admission to critical care; and no COVID-19 deaths. There were 3 non-COVID-19 related deaths; 3,444 fractures; 22,764 A&E attendances; 2,664 unplanned hospitalisations; 9 cases of pericarditis and 3 cases of myocarditis. All pericarditis and myocarditis events occurred in the first dose group, while all COVID-19 related critical care admissions were in the unvaccinated group. Further analyses were restricted to positive SARS-CoV-2 tests, COVID-19 A&E attendance, COVID-19 hospitalisation, fractures, A&E attendance and unplanned hospitalisation.

The incidence of positive SARS-CoV-2 test after first vaccine dose in adolescents declined substantially between 10 days and 6 weeks after vaccination, then increased (Figure 1). By 15 weeks the cumulative incidence of positive SARS-CoV-2 tests was similar in the first dose and unvaccinated groups. The 20-week risks per 10,000 were 1,961 (95% CI 1,932-1,990) and 1,979 (1,950-2,008) in the vaccinated and unvaccinated groups respectively. The IRR comparing vaccinated with unvaccinated adolescents was 0.74 (95% CI 0.72-0.75) (Table 2).

**Figure 1:**
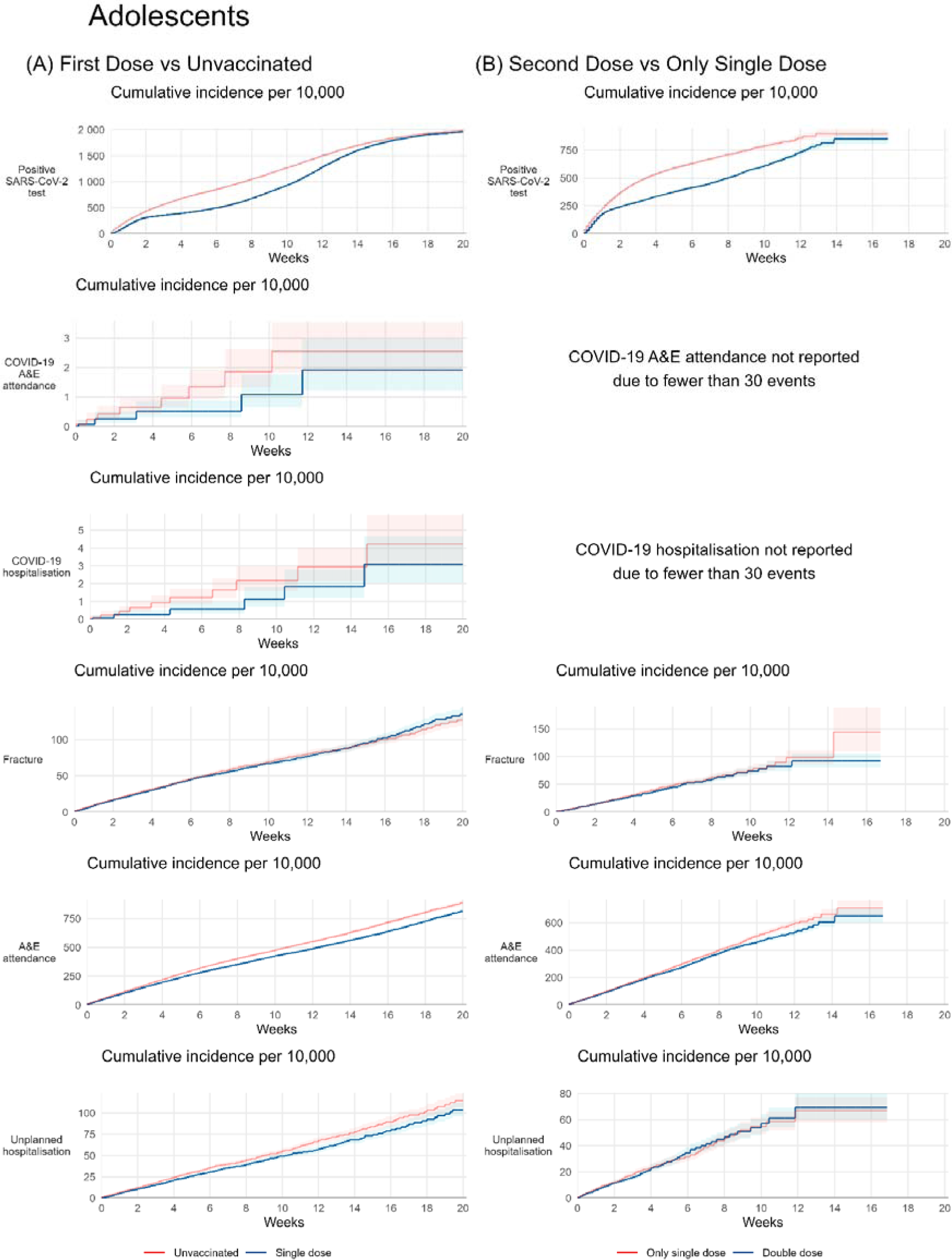
Kaplan-Meier estimates of cumulative incidence and risk ratios of outcomes.

**Figure 2:**
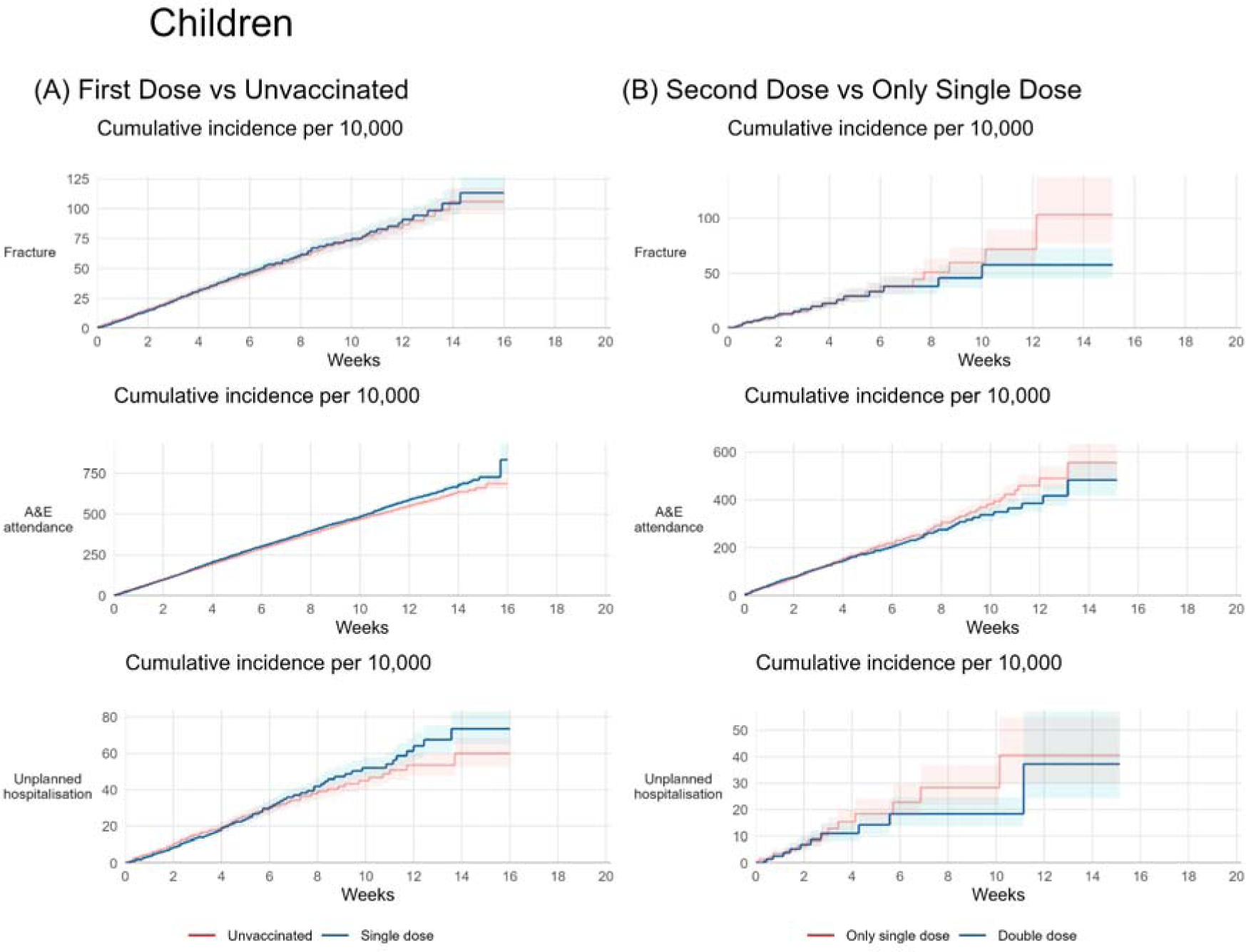
Kaplan-Meier estimates of cumulative incidence and risk ratios of outcomes.

**Table 2:** 20-week risks, events per person years, risk differences and incident rate ratios for adolescents. IRR = Incidence Rate Ratio, Counts and survival estimates are based on values rounded up to the nearest n*6-3, for disclosure control.

| Outcome | Events / Person-years | 20-week risk / 10,000 (95% CI) | Events / Person-years | 20-week risk / 10,000 (95% CI) | RD per 10,000 people (95% CI) | IRR (95% CI) |
| --- | --- | --- | --- | --- | --- | --- |
| First Dose vs Unvaccinated (N = 410,463 per group) |  |  |  |  |  |  |
|  | First Dose |  | Unvaccinated |  |  |  |
| Positive SARS-CoV-2 test | 25,287 / 45,064 | 1,961 (1,932-1,990) | 31,209 / 40,990 | 1,979 (1,950 to 2,008) | -18 (-59 to 23) | 0.74 (0.72 to 0.75) |
| COVID-19 A&E attendance | 27 / 47,801 | 1.91 (1.23 to 2.98) | 45 / 47,827 | 2.54 (1.83 to 3.54) | -0.63 (-1.83 to 0.56) | 0.60 (0.37 to 0.97) |
| COVID-19 hospitalisation | 33 / 47,799 | 3.09 (2.05 to 4.67) | 57 / 47,826 | 4.23 (3.05 to 5.87) | -1.14 (-3.02 to 0.74) | 0.58 (0.38 to 0.89) |
| COVID-19 critical care | 0 / 47,804 | - | 3 / 47,837 | 0.24 (0.08 to 0.76) | -0.24 (-0.52 to 0.03) | - |
| COVID-19 death | 0 / 47,804 | - | 0 / 47,837 | - | - | - |
| Non-COVID-19 death | 3 / 47,804 | 0.51 (0.16 to 1.58) | 0 / 47,837 | - | 0.51 (-0.07 to 1.09) | - |
| Fracture | 1,713 / 47,624 | 135 (126 to 144) | 1,731 / 47,633 | 127 (119 to 136) | 8 (-4 to 20) | 0.99 (0.93 to 1.06) |
| A&E attendance | 10,743 / 46,601 | 812 (792 to 834) | 12,021 / 46,435 | 885 (864 to 907) | -73 (-102 to -43) | 0.89 (0.87 to 0.91) |
| Unplanned hospitalisation | 31,245 / 47,671 | 104 (96 to 112) | 1,419 / 47,656 | 115 (107 to 123) | -11 (-22 to 0) | 0.88 (0.81 to 0.95) |
| Pericarditis events | 9 / 47,802 | 0.31 (0.16 to 0.61) | 0 / 47,837 | - | 0.31 (0.11 to 0.52) | - |
| Myocarditis events | 3 / 47,803 | 0.08 (0.03 to 0.24) | 0 / 47,837 | - | 0.08 (-0.01 to 0.17) | - |
| Second Dose vs Single Dose Only (N = 220,929 per group) |  |  |  |  |  |  |
|  | Second Dose |  | Single Dose Only |  |  |  |
| Positive SARS-CoV-2 test | 6,243 / 14,788 | 850 (802 to 899) | 8,667 / 13,732 | 898 (861 to 935) | -48 (-109 to 13) | 0.67 (0.65 to 0.69) |
| COVID-19 A&E attendance | 3 / 15,220 | 0.14 (0.05 to 0.44) | 3 / 15,223 | 0.14 (0.04 to 0.42) | 0.01 (-0.22 to 0.23) | 1.00 (0.20 to 4.96) |
| COVID-19 hospitalisation | 9 / 15,219 | 0.57 (0.29 to 1.10) | 15 / 15,222 | 2.02 (1.12 to 3.62) | -1.45 (-2.69 to -0.21) | 0.60 (0.26 to 1.37) |
| COVID-19 critical care | 0 / 15,220 | - | 0 / 15,224 | - | - | - |
| COVID-19 death | 0 / 15,220 | - | 0 / 15,224 | - | - | - |
| Non-COVID-19 death | 0 / 15,220 | - | 0 / 15,224 | - | - | - |
| Fracture | 561 / 15,182 | 92 (81 to 105) | 597 / 15,184 | 144 (110 to 189) | -52 (-92 to -11) | 0.94 (0.84 to 1.05) |
| A&E attendance | 3,615 / 14,982 | 651 (602 to 703) | 3,873 / 14,957 | 709 (658 to 763) | -58 (-132 to 15) | 0.93 (0.89 to 0.98) |
| Unplanned hospitalisation | 435 / 15,194 | 69 (60 to 80) | 441 / 15,189 | 67 (58 to 77) | 2 (-11 to 16) | 0.99 (0.86 to 1.13) |
| Pericarditis events | 3 / 15,220 | 0.21 (0.07 to 0.66) | 0 / 15,224 | - | 0.21 (-0.03 to 0.45) | - |
| Myocarditis events | 0 / 15,220 | - | 0 / 15,224 | - | - | - |

**Table 3:** 20-week risks, events per person years, risk differences and incident rate ratios for children. IRR = Incidence Rate Ratio, RD = Risk Difference. Counts and survival estimates are based on values rounded up to the nearest n*6-3, for disclosure control.

| Outcome | Events / Person-years | 20-week risk / 10,000 (95% CI) | Events / Person-years | 20-week risk / 10,000 (95% CI) | RD per 10,000 people (95% CI) | IRR (95% CI) |
| --- | --- | --- | --- | --- | --- | --- |
| First Dose vs Unvaccinated (N = 141,711 per group) |  |  |  |  |  |  |
|  | First Dose |  | Unvaccinated |  |  |  |
| COVID-19 A&E attendance | 0 / 16,233 | - | 0 / 16,243 | - | - | - |
| COVID-19 hospitalisation | 3 / 16,233 | 0.26 (0.08 to 0.79) | 3 / 16,242 | 0.27 (0.09 to 0.85) | -0.02 (-0.44 to 0.41) | 1.00 (0.20 to 4.96) |
| COVID-19 critical care | 0 / 16,233 | - | 0 / 16,243 | - | - | - |
| COVID-19 death | 0 / 16,233 | - | 0 / 16,243 | - | - | - |
| Non-COVID-19 death | 0 / 16,233 | - | 0 / 16,243 | - | - | - |
| Fracture | 633 / 16,175 | 113 (102 to 127) | 621 / 16,183 | 106 (95 to 118) | 7 (-9 to 24) | 1.02 (0.91 to 1.14) |
| A&E attendance | 4,107 / 15,862 | 834 (738 to 929) | 3,909 / 15,878 | 687 (653 to 723) | 146 (49 to 244) | 1.05 (1.01 to 1.10) |
| Unplanned hospitalisation | 417 / 16,196 | 73 (65 to 83) | 381 / 16,204 | 60 (52 to 69) | 14 (1 to 26) | 1.10 (0.95 to 1.26) |
| Pericarditis events | 3 / 16,233 | 0.22 (0.07 to 0.67) | 0 / 16,243 | - | 0.22 (-0.03 to 0.46) | - |
| Myocarditis events | 0 / 16,233 | - | 0 / 16,243 | - | - | - |
| Second Dose vs Single Dose Only (N = 66,231 per group) |  |  |  |  |  |  |
|  | Second Dose |  | Single Dose Only |  |  |  |
| COVID-19 A&E attendance | 0 / 4,436 | - | 0 / 4,439 | - | - | - |
| COVID-19 hospitalisation | 0 / 4,436 | - | 0 / 4,439 | - | - | - |
| COVID-19 critical care | 0 / 4,436 | - | 0 / 4,439 | - | - | - |
| COVID-19 death | 0 / 4,436 | - | 0 / 4,439 | - | - | - |
| Non-COVID-19 death | 0 / 4,436 | - | 0 / 4,439 | - | - | - |
| Fracture | 123 / 4,427 | 58 (45 to 73) | 141 / 4,431 | 103 (77 to 138) | -46 (-78 to -13) | 0.87 (0.69 to 1.11) |
| A&E attendance | 819 / 4,387 | 483 (419 to 557) | 861 / 4,382 | 556 (488 to 632) | -73 (-172 to 27) | 0.95 (0.86 to 1.05) |
| Unplanned hospitalisation | 63 / 4,432 | 37 (24 to 57) | 81 / 4,431 | 41 (30 to 55) | -3 (-24 to 17) | 0.78 (0.56 to 1.08) |
| Pericarditis events | 0 / 4,436 | - | 0 / 4,439 | - | - | - |
| Myocarditis events | 0 / 4,436 | - | 0 / 4,439 | - | - | - |

The incidence of COVID-19 A&E attendance was lower after first vaccination than in the unvaccinated group (IRR 0.60; 95% CI 0.37-0.97). The 20-week risks per 10,000 were 1.91 (95% CI 1.23-2.98) and 2.54 (1.83-3.54) respectively. The incidence of COVID-19 hospitalisation was lower after first vaccination than in the unvaccinated group (IRR 0.58; 0.38-0.89). The 20-week risks per 10,000 were 3.09 (2.05-4.67) and 4.23 (3.05-5.87) respectively.

The incidence of fractures (negative control outcome) was similar in the first vaccination and unvaccinated groups: (IRR 0.99; 95% CI 0.93-1.06). The 20-week risks per 10,000 were 135 (95% CI 126-144) and 127 (119-136) respectively. The incidence of A&E attendance (safety outcome) was lower after first vaccination than in the unvaccinated group (IRR 0.89; 0.87-0.91). The 20-week risks per 10,000 were 812 (95% CI 792-834) and 885 (864-907) respectively. The incidence of unplanned hospitalisation was lower after first vaccination than in the unvaccinated group (IRR 0.88; 0.81-0.95). The 20-week risks per 10,000 were 104 (96-112) and 115 (107-123) respectively.

### Effectiveness of second dose vs single dose only in adolescents

In 30,444 person-years follow-up after first vaccination in adolescents, there were 14,910 positive SARS-CoV-2 tests; 6 COVID-19 A&E attendances; 24 COVID-19 hospitalisations, none of which included admission to critical care; and no COVID-19 deaths. There were no non-COVID-19 related deaths; 1,158 fractures; 7,488 A&E attendances; 876 unplanned hospitalisations; 3 cases of pericarditis and no cases of myocarditis. All pericarditis events occurred in the single dose group. Further analyses were restricted to positive SARS-CoV-2 tests, A&E attendance, fractures and unplanned hospitalisation.

The incidence of positive SARS-CoV-2 test after second dose declined between 10 days and 6 weeks after vaccination, then increased (Figure 1). By 14 weeks the cumulative incidence of positive SARS-CoV-2 test was similar in the second and single dose groups. The 20-week risks per 10,000 were 850 (95% CI 802-899) and 898 (861-935) after second and single dose respectively. The IRR comparing second with single doses was 0.67 (95% CI 0.65-0.69) (Table 2).

The IRR comparing the incidence of fractures (negative control outcome) was similar in the second and single dose groups was 0.94 (95% CI 0.84-1.05). The 20-week risks per 10,000 were 92 (95% CI 81-105) and 144 (110-189) respectively. The incidence of A&E attendance (safety outcome) was lower after second than single dose (IRR 0.93; 0.89-0.98). The 20-week risks per 10,000 were 651 (95% CI 602-703) and 709 (658-763) respectively. The incidence of unplanned hospitalisation was similar in the second and single dose groups (IRR 0.99; 0.86-1.13). The 20-week risks per 10,000 were 69 (60-80) and 67 (58-77) respectively.

### Myocarditis and pericarditis in adolescents

Less than 51% of adolescents diagnosed with pericarditis were admitted to hospital, and less than 51% attended emergency care. More than 51% of adolescents with myocarditis were admitted to hospital and more than 51% attended emergency care. The maximum length of critical care admission was 1 day for either event. The maximum length of hospitalisation stay was 0 days for pericarditis and 2 days for myocarditis. There were no deaths after these events.

### Children

Of 177,360 children eligible in the first vaccination group 141,711 (80%) were matched with unvaccinated controls. (Supplementary Figure 2, panel A). 66,231 (67%) of 99,102 children who received a second BNT162b2 vaccination were matched with single vaccinated control. (Supplementary Figure 2, panel 2). Nearly 60% of children in the first vaccination group received a second vaccination. Most second vaccinations occurred at least 12 weeks after the first vaccination (Supplementary Figure 2).

### Effectiveness of first dose vs unvaccinated in children

In 32,476 person-years follow-up, there were no COVID-19 A&E attendances; 6 COVID-19 hospitalisations (none included admission to critical care) and no COVID-19 or non-COVID-19 related deaths. There were 1,254 fractures; 8,016 A&E attendances; 798 unplanned hospitalisations; 3 cases of pericarditis and no cases of myocarditis. All pericarditis events occurred in the first dose group. Further analyses were restricted to fractures, A&E attendance and unplanned hospitalisation. The incidence of fractures was similar in the first vaccination and unvaccinated groups (IRR 1.02; 95% CI 0.91-1.14). The 20-week risks per 10,000 were 113 (95% CI 102-127) and 106 (95-118) respectively. The incidence of A&E attendance was higher after first vaccination than in the unvaccinated group (IRR 1.05; 95% CI 1.01-1.10). The 20-week risks per 10,000 were 834 (748-929) and 687 (653-723) respectively. The IRR comparing the incidence of unplanned hospitalisation in the first vaccination and unvaccinated groups was 1.10 (95% CI 0.95-1.26). The 20-week risks per 10,000 were 73 (65-83) and 60 (52-69) respectively.

### Effectiveness of second dose vs single dose only in children

In 8,875 person-years follow-up, there were no COVID-19 A&E attendances; no COVID-19 hospitalisations; no admissions to critical care; and no COVID-19 or non-COVID-19 related deaths. There were 264 fractures; 1,680 A&E attendances; 144 unplanned hospitalisations; and no cases of pericarditis or myocarditis. Further analyses were restricted to fractures, A&E attendance and unplanned hospitalisation.

The IRR comparing the incidence of fractures in the second and single dose groups was 0.87 (95% CI 0.69-1.11). The 20-week risks per 10,000 were 58 (95% CI 45-73) and 103 (77-138) respectively. The incidence of A&E attendance was similar in the second and single dose groups (IRR 0.95; 0.86-1.05). The 20-week risks per 10,000 were 483 (419-557) and 556 (488-632) respectively. The IRR comparing the incidence of unplanned hospitalisation in the second and single dose groups was 0.78 (95% CI 0.56-1.08). The 20-week risks per 10,000 were 37 (24-57) and 41 (30-55) respectively.

### Myocarditis and pericarditis events in children

No children experienced a myocarditis event, all 3 pericarditis events occurred after first vaccination and did not require hospitalisation or critical care.

## Discussion

This observational cohort study of COVID-19 vaccination with BNT162b2 in England based on 410,463 adolescents (12-15 years old) receiving a first vaccination, 220,029 adolescents receiving a second vaccination, 141,711 children (5-11 years old) receiving a first vaccination and 66,231 children receiving a second vaccination, found that an initial protective effect against positive SARS-CoV-2 test in adolescents waned by 14 weeks. The incidence of COVID-19 A&E attendance was lower after first vaccination than in the unvaccinated group for adolescents and COVID-19 A&E attendance was rare in the other adolescent group and the child groups. COVID-19-related hospitalisation, and critical care attendance were rare in both adolescents and children and there were no COVID-19 related deaths. Whilst rare, all myocarditis and pericarditis events during the study period occurred in vaccinated individuals: there were no deaths after myocarditis or pericarditis. The rate of fractures was similar across vaccine groups in both adolescents and children. None of the child cohort required hospitalisation or critical care after a pericarditis event. In the adolescents the maximum length of hospital admission was 1 day for critical care and 2 days for hospitalisation. Our findings provide insights into the balance between protection by vaccination against target outcomes (positive SARS-CoV-2 tests, COVID-19 related hospitalisation and emergency care) and the increased risk of pericarditis and myocarditis. In adolescents the reduction in risk of COVID-19 hospitalisation per 10,000 individuals (-1.14 for first dose versus unvaccinated, -1.45 for second versus first dose) was larger than the increase in risk of both myocarditis (0.08 for first dose versus unvaccinated) and pericarditis (0.31 for first dose versus unvaccinated, 0.21 for second versus first dose). However, the reduction in risk of COVID-19 hospitalisation in children (-0.02 for first dose versus unvaccinated) was lower than the increase in risk of pericarditis (0.22).

### Strengths and Limitations

Our study has several limitations. First, bias due to unmeasured confounding is possible, although the detailed data available in OpenSAFELY allowed us to match vaccinated with control individuals on multiple characteristics. For example, it was not possible to reliably identify individuals with COVID-19 symptoms prior to testing or hospital admission, because this information is not routinely recorded in primary care records. Delayed vaccination in individuals with symptoms may explain the apparent effectiveness against positive tests during the first week, however, the incidence of positive tests substantially reduced from 1-2 weeks in the vaccinated group (Figure 1A) compared with the unvaccinated group, consistent with a delayed but protective immunological response to vaccination. A similar pattern was observed for second dose versus single dose vaccination (Figure 1B). Lifestyle factors that may influence both health and vaccination are not fully recorded in health records. The slightly higher rate of fractures (negative control outcome) in single-vaccinated adolescents and children compared to the second dose group could indicate limited unmeasured confounding. Second, positive SARS-CoV-2 test data underestimates the true incidence of infection. Both lateral flow tests and PCR tests were freely available in the UK until 31 March 2023, but many asymptomatic and some symptomatic infections will not have been recorded. Differences in testing behaviour between vaccinated and unvaccinated people may have contributed to the apparent waning effect if testing is more common among vaccinated individuals. Third, myocarditis and pericarditis following COVID-19 vaccination were publicly reported from May 2021(12). The vaccinations in our study were after this date, so there is a potential for ascertainment bias if diagnostic thresholds were lower in vaccinated than unvaccinated individuals. Fourth, we excluded clinically vulnerable children and adolescents who were eligible for vaccination before the general roll out, so our results may not be generalisable to this group. Vaccine safety and effectiveness in this group are important, but there are significant challenges in controlling confounding arising from unmeasured severity of underlying conditions that could influence vaccine uptake and effectiveness. Fifth, due to small numbers we did not study vaccine effectiveness in those who had received a vaccination other than BNT162b2.

### Findings in context

BNT162b2 vaccination was shown to be effective in protecting against COVID-19 infection in a multinational phase 3 trial of 2,260 adolescents aged 12-15 with a median follow up of 2 months(13,14). Multiple observational studies have found that effectiveness wanes with time since vaccination(15,16). A recent systematic review found generally limited evidence regarding clinical outcomes of BNT162b2 vaccination amongst children and adolescents(17), although vaccination lowered hospitalisation rates including emergency admission.

Several studies have reported that children infected with SARS-CoV-2 appear to have the same degree of severity of illness as seen in adults(18–20). Most reported post-vaccination cases of myocarditis and pericarditis in children and adolescents were mild(21). A systematic review found that myocarditis and pericarditis in children and adolescents were more prevalent among males and after the second dose of BNT162b2(22). Whilst all cases of myocarditis and pericarditis in our study occurred in the vaccinated groups, we did not find higher rates of myocarditis or pericarditis after the second compared to the first dose. The rates of heart inflammation (myocarditis and pericarditis) in under 18s reported by the UK Medicines and Healthcare products Regulatory Agency were 13 and 8 per million after first and second doses respectively, compared with our estimates of 27 and 10 per million respectively.

## Conclusion

This study found that initial protection by BNT162b2 vaccination against positive SARS-CoV-2 tests in adolescents aged 12-15 had waned by 14 weeks after vaccination. Rates of COVID-19 hospitalisation and COVID-19 A&E attendance were lower after first and second dose BNT162b2 vaccination in adolescents. Positive SARS-CoV-2 testing could not be considered for children. Severe outcomes were rare in children: there were fewer than seven (exact number redacted) COVID-19 hospitalisations and no COVID-19 A&E attendances, critical care admissions or COVID-19 deaths.

## Administrative

## Acknowledgements

We are very grateful for all the support received from the TPP Technical Operations team throughout this work, and for generous assistance from the information governance and database teams at NHS England and the NHS England Transformation Directorate

## Information governance and ethical approval

NHS England is the data controller of the NHS England OpenSAFELY COVID-19 Service; TPP is the data processor; all study authors using OpenSAFELY have the approval of NHS England(22). This implementation of OpenSAFELY is hosted within the TPP environment which is accredited to the ISO 27001 information security standard and is NHS IG Toolkit compliant(23).

Patient data has been pseudonymised for analysis and linkage using industry standard cryptographic hashing techniques; all pseudonymised datasets transmitted for linkage onto OpenSAFELY are encrypted; access to the NHS England OpenSAFELY COVID-19 service is via a virtual private network (VPN) connection; the researchers hold contracts with NHS England and only access the platform to initiate database queries and statistical models; all database activity is logged; only aggregate statistical outputs leave the platform environment following best practice for anonymisation of results such as statistical disclosure control for low cell counts(24).

The service adheres to the obligations of the UK General Data Protection Regulation (UK GDPR) and the Data Protection Act 2018. The service previously operated under notices initially issued in February 2020 by the the Secretary of State under Regulation 3(4) of the Health Service (Control of Patient Information) Regulations 2002 (COPI Regulations), which required organisations to process confidential patient information for COVID-19 purposes; this set aside the requirement for patient consent(25). As of 1 July 2023, the Secretary of State has requested that NHS England continue to operate the Service under the COVID-19 Directions 2020(26). In some cases of data sharing, the common law duty of confidence is met using, for example, patient consent or support from the Health Research Authority Confidentiality Advisory Group(27).

Taken together, these provide the legal bases to link patient datasets using the service. GP practices, which provide access to the primary care data, are required to share relevant health information to support the public health response to the pandemic, and have been informed of how the service operates.

This study was approved by the Health Research Authority (REC reference 20/LO/0651) and by the London School of Hygeine and Tropical Medicine Ethics Board (reference 21863).

## Guarantor

WHJ is guarantor

## Supplementary

**Supplementary Table 1:** Adolescents First Dose vs Unvaccinated subgroup analyses according to prior infection/COVID.

| First Dose vs Unvaccinated |  |  |  |  |  |  |  |  |
| --- | --- | --- | --- | --- | --- | --- | --- | --- |
| Outcome | Sub-group | N per group | Unvaccinated |  | First Dose |  | RD per 10,000 people (95% CI) | IRR (95% CI) |
|  |  |  | Events / Person-years | 20-week risk / 10,000 (95% CI) | Events / Person-years | 20-week risk / 10,000 people (95% CI) |  |  |
| Positive SARS-CoV-2 test | No prior SARS-CoV-2 infection | 328,323 | 29,223 / 31,024 | 2,255 (2,221 to 2,289) | 23,943 / 34,851 | 2,263 (2,221 to 2,289) | 8 (–39 to 56) | 0.73 (0.72 to 0.74) |
|  | Prior SARS-CoV-2 infection | 82,137 | 1,983 / 9,965 | 1,029 (974 to 1,087) | 1,341 / 10,212 | 717 (974 to 1,087) | –313 (–385 to –240) | 0.66 (0.62 to 0.71) |
| COVID-19 A&E attendance | No prior SARS-CoV-2 infection | 328,323 | 45 / 37,486 | 4.16 (2.86 to 6.03) | 21 / 37,466 | 1.54 (0.95 to 2.51) | –2.61 (–4.33 to –0.89) | 0.47 (0.28 to 0.78) |
|  | Prior SARS-CoV-2 infection | 82,137 | 3 / 10,340 | 0.76 (0.24 to 2.35) | 3 / 10,334 | 0.47 (0.15 to 1.45) | –0.29 (–1.30 to 0.72) | 1.00 (0.20 to 4.96) |
| COVID-19 hospitalisation | No prior SARS-CoV-2 infection | 328,323 | 51 / 37,484 | 4.04 (2.93 to 5.57) | 33 / 37,465 | 5.46 (3.44 to 8.66) | 1.42 (–1.41 to 4.25) | 0.65 (0.42 to 1.00) |
|  | Prior SARS-CoV-2 infection | 82,137 | 3 / 10,340 | 0.53 (0.17 to 1.63) | 3 / 10,334 | 0.54 (0.18 to 1.68) | 0.02 (–0.84 to 0.87) | 1.00 (0.20 to 4.96) |
| Fracture | No prior SARS-CoV-2 infection | 328,323 | 1,335 / 37,334 | 124 (115 to 133) | 1,311 / 37,328 | 131 (115 to 133) | 7 (–6 to 20) | 0.98 (0.91 to 1.06) |
|  | Prior SARS-CoV-2 infection | 82,137 | 399 / 10,298 | 150 (128 to 175) | 399 / 10,296 | 149 (128 to 175) | –1 (–33 to 30) | 1.00 (0.87 to 1.15) |
| A&E attendance | No prior SARS-CoV-2 infection | 328,323 | 9,219 / 36,404 | 864 (841 to 887) | 8,097 / 36,553 | 780 (841 to 887) | –83 (–116 to –51) | 0.87 (0.85 to 0.90) |
|  | Prior SARS-CoV-2 infection | 82,137 | 2,799 / 10,030 | 977 (925 to 1,032) | 2,643 / 10,047 | 945 (925 to 1,032) | –33 (–108 to 42) | 0.94 (0.89 to 0.99) |
| Unplanned hospitalisation | No prior SARS-CoV-2 infection | 328,323 | 1,095 / 37,355 | 114 (105 to 123) | 939 / 37,367 | 100 (105 to 123) | –14 (–27 to –2) | 0.86 (0.79 to 0.94) |
|  | Prior SARS-CoV-2 infection | 82,137 | 327 / 10,300 | 126 (107 to 148) | 309 / 10,303 | 130 (107 to 148) | 4 (–25 to 33) | 0.94 (0.81 to 1.10) |

20-week risks, events per person years, risk differences and incident rate ratios for adolescents, first vaccination). Counts and survival estimates are based on values rounded up to the nearest n\*6-3, for disclosure control. IRR Incidence Rate Ratio, RD = Risk Difference, RR = Risk Ratio, (95% Confidence Intervals). Counts and survival estimates are based on values rounded up to the nearest n\*6-3, for disclosure control.

**Supplementary Table 2:** Adolescents Second Dose vs Single Dose Only subgroup analyses according to prior infection/COVID.

| Second Dose vs Single Dose Only |  |  |  |  |  |  |  |  |
| --- | --- | --- | --- | --- | --- | --- | --- | --- |
| Outcome | Sub-group | N per group | Single Dose Only |  | Second Dose |  | RD per 10,000 people (95% CI) | IRR (95% CI) |
|  |  |  | Events / Person-years | 20-week risk / 10,000 people (95% CI) | Events / Person-years | 20-week risk / 10,000 people (95% CI) |  |  |
| Positive SARS-CoV-2 test | No prior SARS-CoV-2 infection | 170,919 | 7,827 / 10,317 | 1,061 (1,012 to 1,112) | 5,661 / 11,286 | 1,144 (1,012 to 1,112) | 83 (−88 to 253) | 0.66 (0.64 to 0.68) |
|  | Prior SARS-CoV-2 infection | 50,007 | 843 / 3,414 | 449 (395 to 510) | 585 / 3,501 | 341 (395 to 510) | −108 (−184 to −31) | 0.68 (0.61 to 0.75) |
| COVID-19 A&E attendance | No prior SARS-CoV-2 infection | 170,919 | 3 / 11,687 | 0.18 (0.06 to 0.54) | 3 / 11,684 | 0.19 (0.06 to 0.57) | 0.01 (−0.28 to 0.30) | 1.00 (0.20 to 4.96) |
|  | Prior SARS-CoV-2 infection | 50,007 | 0 / 3,536 | - | 0 / 3,535 | - | - | - |
| COVID-19 hospitalisation | No prior SARS-CoV-2 infection | 170,919 | 9 / 11,686 | 0.95 (0.48 to 1.87) | 9 / 11,684 | 0.83 (0.42 to 1.62) | −0.12 (−0.97 to 0.73) | 1.00 (0.40 to 2.52) |
|  | Prior SARS-CoV-2 infection | 50,007 | 3 / 3,535 | 0.60 (0.19 to 1.86) | 3 / 3,535 | 0.70 (0.23 to 2.18) | 0.10 (−0.94 to 1.15) | 1.00 (0.20 to 4.96) |
| Fracture | No prior SARS-CoV-2 infection | 170,919 | 435 / 11,657 | 121 (98 to 147) | 417 / 11,655 | 97 (98 to 147) | −23 (−52 to 6) | 0.96 (0.84 to 1.10) |
|  | Prior SARS-CoV-2 infection | 50,007 | 159 / 3,526 | 81 (66 to 101) | 147 / 3,526 | 105 (66 to 101) | 23 (−11 to 58) | 0.92 (0.74 to 1.16) |
| A&E attendance | No prior SARS-CoV-2 infection | 170,919 | 2,793 / 11,490 | 685 (634 to 740) | 2,619 / 11,506 | 654 (634 to 740) | −31 (−122 to 61) | 0.94 (0.89 to 0.99) |
|  | Prior SARS-CoV-2 infection | 50,007 | 1,077 / 3,466 | 679 (609 to 758) | 999 / 3,476 | 1,007 (609 to 758) | 327 (71 to 584) | 0.93 (0.85 to 1.01) |
| Unplanned hospitalisation | No prior SARS-CoV-2 infection | 170,919 | 315 / 11,662 | 59 (51 to 68) | 315 / 11,665 | 66 (51 to 68) | 7 (−6 to 20) | 1.00 (0.86 to 1.17) |
|  | Prior SARS-CoV-2 infection | 50,007 | 123 / 3,526 | 66 (52 to 84) | 117 / 3,528 | 61 (52 to 84) | −5 (−26 to 15) | 0.95 (0.74 to 1.22) |

**Supplementary Table 3:** Children First Dose vs Unvaccinated subgroup analyses according to prior infection/COVID.

| Outcome | Sub-group | N per group | First Dose vs Unvaccinated |  |  |  |  | IRR (95% CI) |
| --- | --- | --- | --- | --- | --- | --- | --- | --- |
|  |  |  | Unvaccinated |  | First Dose |  | RD per 10,000 people (95% CI) |  |
|  |  |  | Events / Person-years | 20-week risk per 10,000 people (95% CI) | Events / Person-years | 20-week risk per 10,000 people (95% CI) |  |  |
| COVID-19 hospitalisation | No prior SARS-CoV-2 infection | 76,407 | 3 / 8,759 | 0.69 (0.22 to 2.14) | 3 / 8,751 | 0.48 (0.15 to 1.49) | -0.21 (-1.16 to 0.74) | 1.00 (0.20 to 4.96) |
|  | Prior SARS-CoV-2 infection | 65,307 | 3 / 7,485 | 0.59 (0.19 to 1.82) | 0 / 7,482 | - | -0.59 (-1.25 to 0.08) | - |
| Fracture | No prior SARS-CoV-2 infection | 76,407 | 309 / 8,728 | 96 (83 to 112) | 309 / 8,723 | 104 (83 to 112) | 7 (-15 to 30) | 1.00 (0.85 to 1.17) |
|  | Prior SARS-CoV-2 infection | 65,307 | 315 / 7,457 | 137 (113 to 166) | 327 / 7,453 | 139 (113 to 166) | 2 (-34 to 38) | 1.04 (0.89 to 1.21) |
| A&E attendance | No prior SARS-CoV-2 infection | 76,407 | 1,959 / 8,576 | 658 (610 to 710) | 2,061 / 8,562 | 1,094 (610 to 710) | 436 (112 to 759) | 1.05 (0.99 to 1.12) |
|  | Prior SARS-CoV-2 infection | 65,307 | 1,947 / 7,303 | 704 (661 to 750) | 2,049 / 7,301 | 850 (661 to 750) | 146 (52 to 240) | 1.05 (0.99 to 1.12) |
| Unplanned hospitalisation | No prior SARS-CoV-2 infection | 76,407 | 201 / 8,738 | 55 (47 to 64) | 207 / 8,732 | 63 (47 to 64) | 8 (-5 to 20) | 1.03 (0.85 to 1.25) |
|  | Prior SARS-CoV-2 infection | 65,307 | 177 / 7,467 | 55 (46 to 66) | 207 / 7,465 | 77 (46 to 66) | 22 (6 to 38) | 1.17 (0.96 to 1.43) |
| Pericarditis events | No prior SARS-CoV-2 infection | 76,407 | 0 / 8,759 | - | 0 / 8,752 | - | - | - |
|  | Prior SARS-CoV-2 infection | 65,307 | 0 / 7,485 | - | 3 / 7,482 | 0.47 (0.15 to 1.46) | 0.47 (-0.06 to 1.00) | - |

20-week risks, events per person years, risk differences and incident rate ratios for children first vaccination. Counts and survival estimates are based on values rounded up to the nearest n\*6-3, for disclosure control. IRR Incidence Rate Ratio, RD = Risk Difference, RR = Risk Ratio, (95% Confidence Intervals). Counts and survival estimates are based on values rounded up to the nearest n\*6-3, for disclosure control.

**Supplementary Table 4:** Children Second Dose vs Single Dose Only subgroup analyses according to prior infection/COVID.

| Second Dose vs Single Dose Only |  |  |  |  |  |  |  |  |
| --- | --- | --- | --- | --- | --- | --- | --- | --- |
| Outcome |  | N per group | Single Dose Only |  | Second Dose |  | RD per 10,000 people (95% CI) | IRR (95% CI) |
|  |  |  | Events / Person-years | 20-week risk per 10,000 people (95% CI) | Events / Person-years | 20-week risk per 10,000 people (95% CI) |  |  |
| Fracture | No prior SARS-CoV-2 infection | 35,451 | 63 / 2,431 | 73 (53 to 103) | 57 / 2,428 | 43 (53 to 103) | -31 (-59 to -3) | 0.91 (0.63 to 1.30) |
|  | Prior SARS-CoV-2 infection | 30,777 | 75 / 1,999 | 84 (63 to 113) | 63 / 1,998 | 59 (63 to 113) | -25 (-56 to 6) | 0.84 (0.60 to 1.17) |
| A&E attendance | No prior SARS-CoV-2 infection | 35,451 | 447 / 2,405 | 554 (459 to 669) | 423 / 2,407 | 531 (459 to 669) | -24 (-175 to 128) | 0.95 (0.83 to 1.08) |
|  | Prior SARS-CoV-2 infection | 30,777 | 417 / 1,976 | 969 (666 to 1,400) | 399 / 1,980 | 547 (666 to 1,400) | -422 (-808 to -36) | 0.96 (0.83 to 1.10) |
| Unplanned hospitalisation | No prior SARS-CoV-2 infection | 35,451 | 45 / 2,431 | 82 (48 to 140) | 27 / 2,431 | 15 (48 to 140) | -67 (-111 to -23) | 0.60 (0.37 to 0.97) |
|  | Prior SARS-CoV-2 infection | 30,777 | 39 / 1,999 | 37 (25 to 55) | 33 / 2,001 | 63 (25 to 55) | 26 (-14 to 65) | 0.85 (0.53 to 1.34) |

20-week event counts, Kaplan-Meier risk estimates, and comparisons (children, first vaccination). Counts and survival estimates are based on values rounded up to the nearest n\*6-3, for disclosure control. IRR Incidence Rate Ratio, RD = Risk Difference, RR = Risk Ratio, (95% Confidence Intervals). Counts and survival estimates are based on values rounded up to the nearest n\*6-3, for disclosure control.

**Supplementary Figure 1:**
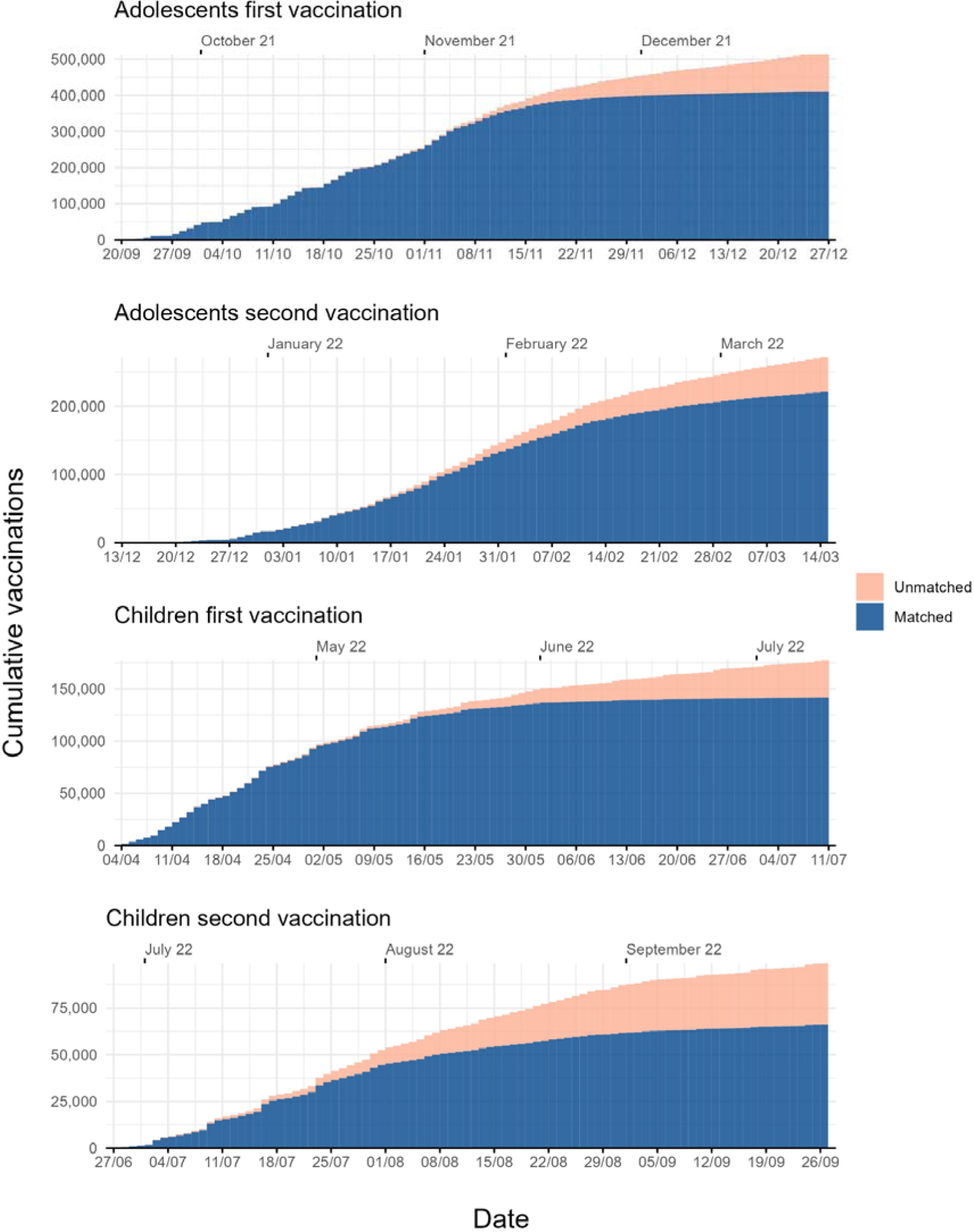
Cumulative matching coverage.

**Supplementary Figure 2:**
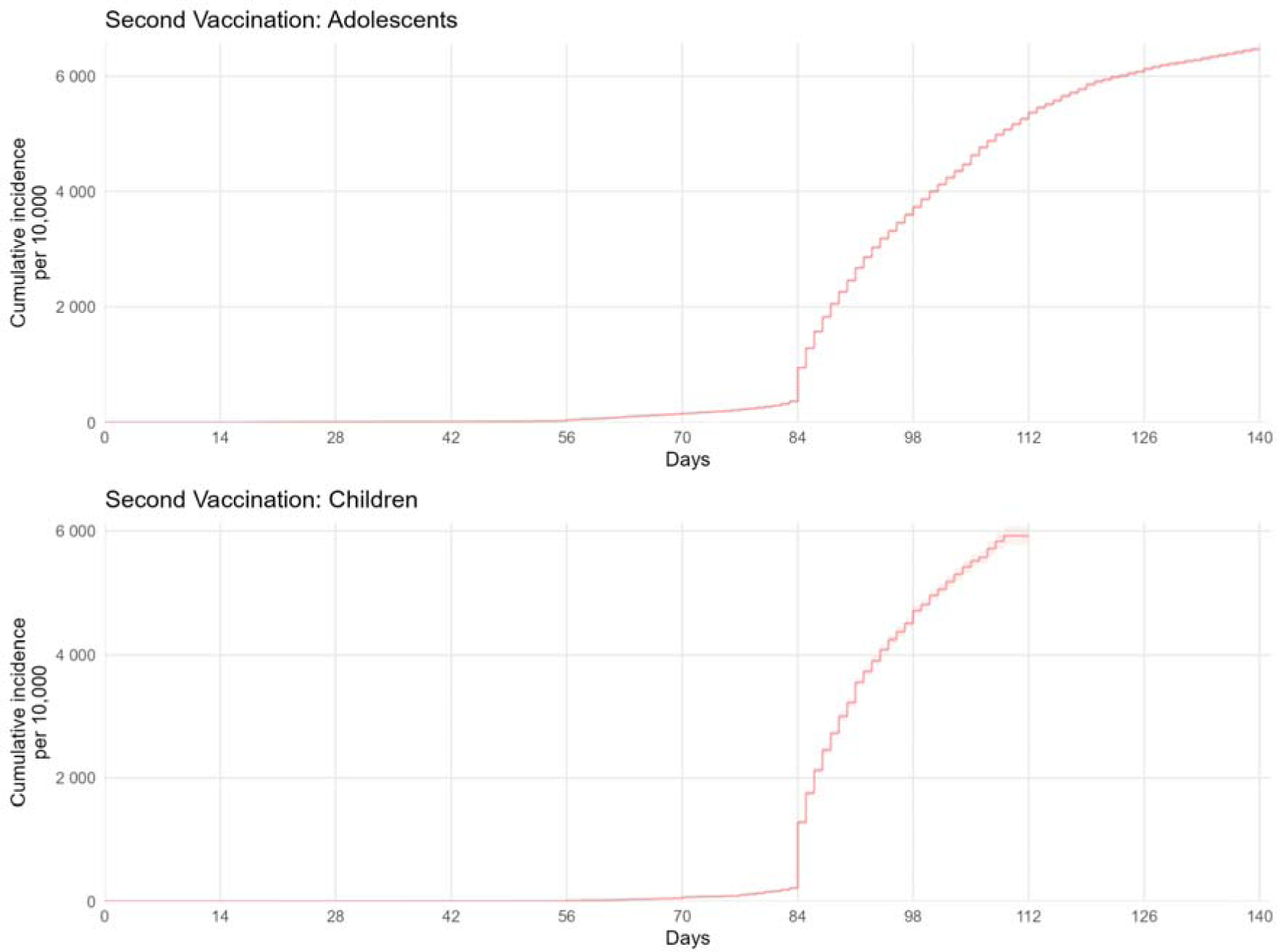
Kaplan-Meier estimates of cumulative incidence of second vaccination for adolescents and children.

**Supplementary Figure 3:**
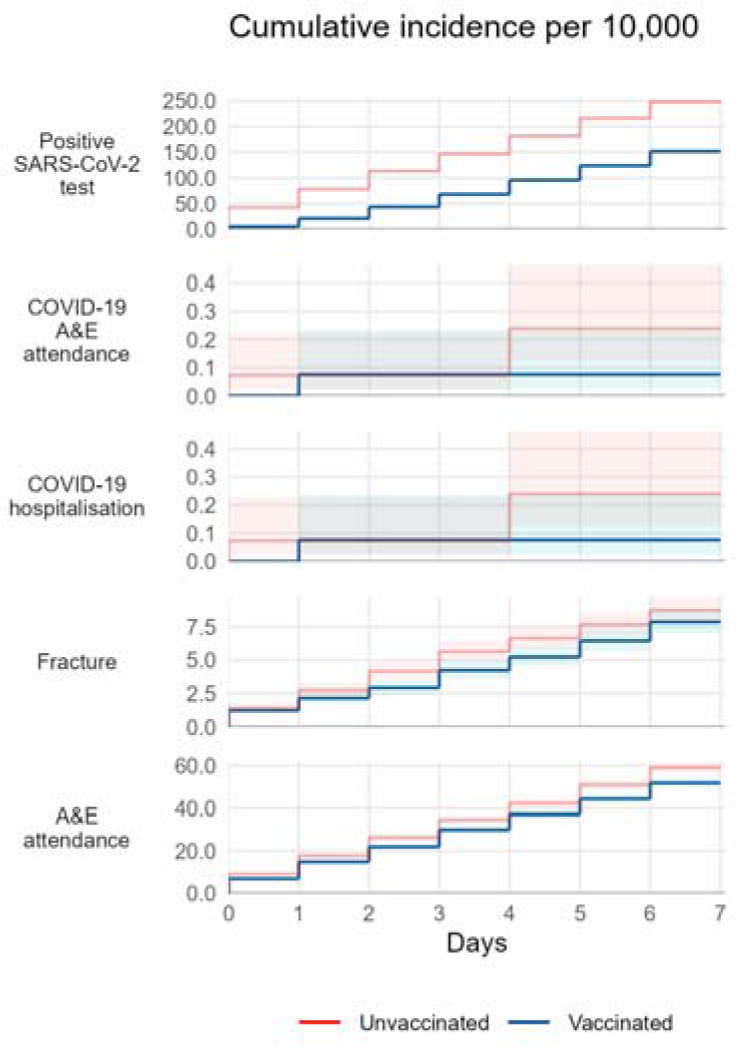
Early outcome Kaplan-Meier estimates of cumulative incidence and risk ratios of outcomes in matched vaccinated and unvaccinated adolescents for the first 7 days.

**Supplementary Figure 4:**
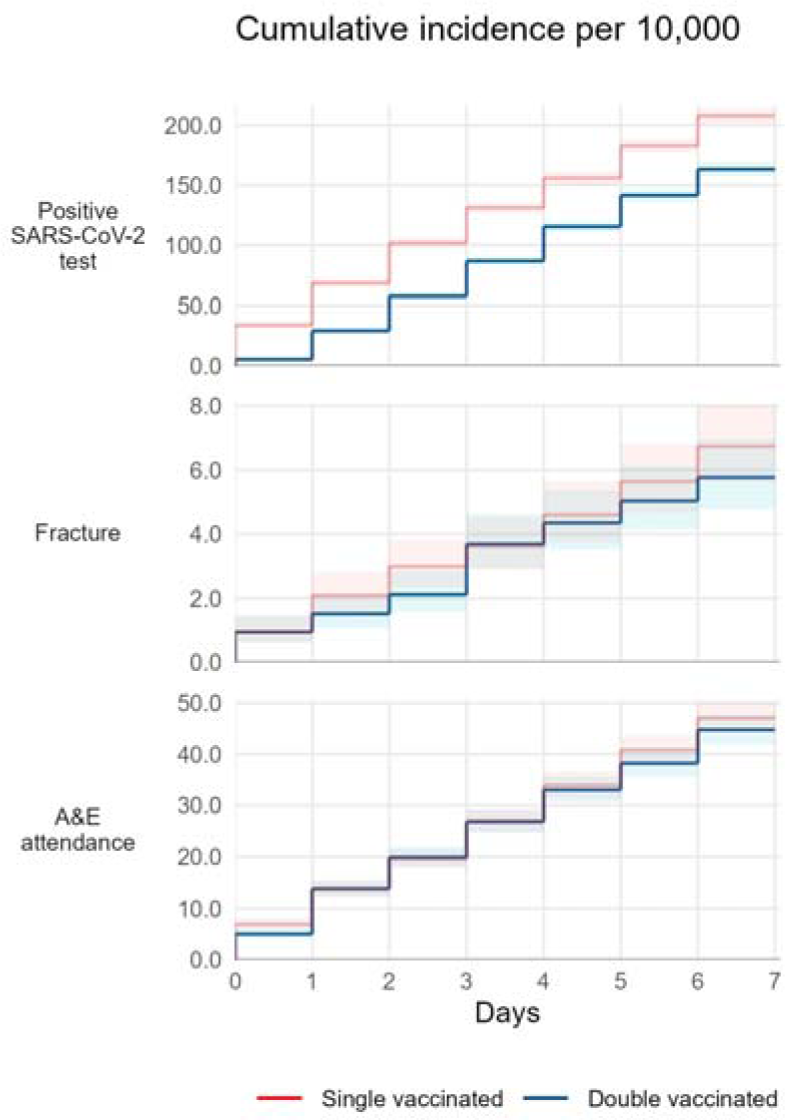
Early outcome Kaplan-Meier estimates of cumulative incidence and risk ratios of outcomes in matched second vaccinated and single vaccinated adolescents for the first 7 days.

**Supplementary Box 1:** Definitions of study outcomes.

| Outcome | Definition |
| --- | --- |
| Positive SARS-CoV-2 test (Adolescents only) | Identified using SGSS testing records and based on swab date. Both polymerase chain reaction (PCR) and lateral flow test results are included, without differentiation between symptomatic and asymptomatic infection. |
| COVID-19 A&E attendances | Identified using HES emergency care records with U07.1 ("COVID-19, virus identified") or U07.2 ("COVID-19, virus not identified") ICD-10 diagnosis codes. |
| COVID-19 related hospital admission, | Identified using HES in-patient hospital records with U07.1 or U07.2 ICD-10 reason for admission codes. |
| COVID-19 critical care admission. | Hospital admissions involving a stay of at least one day on an ICU/HDU/critical care ward. |
| COVID-19 death, | Identified using death records with U07.1 or U07.2 ICD-10 codes present (i.e., as an underlying or contributing cause of death). |
| Non-COVID-19 death | negative control outcomes<sup>11</sup> |
| Fracture | negative control outcomes<sup>11</sup> |
| A&E attendances | Any recorded attendance of A&E from the Emergency Care Data Set (ECDS) during the study period. |
| Unplanned hospitalisation | Identified using HES in-patient hospital records |
| Pericarditis | Identified as either A&E attendances identified using HES emergency care records with 3238004 ("Pericarditis") or 373945007 ("Pericardial effusion") SNOMED diagnosis codes or hospital admission, identified using HES in-patient hospital records with I30 ("Acute pericarditis") ICD-10 reason for admission codes. |
| Myocarditis | Identified as either A&E attendances identified using HES emergency care records with 50920009 ("myocarditis") SNOMED diagnosis codes or hospital admission, identified using HES in-patient hospital records with I514 ("myocarditis, unspecified"), I41 ("myocarditis in diseases classified elsewhere"), or I40 ("Acute myocarditis") ICD-10 reason for admission codes. |

**Supplementary Box 2:**
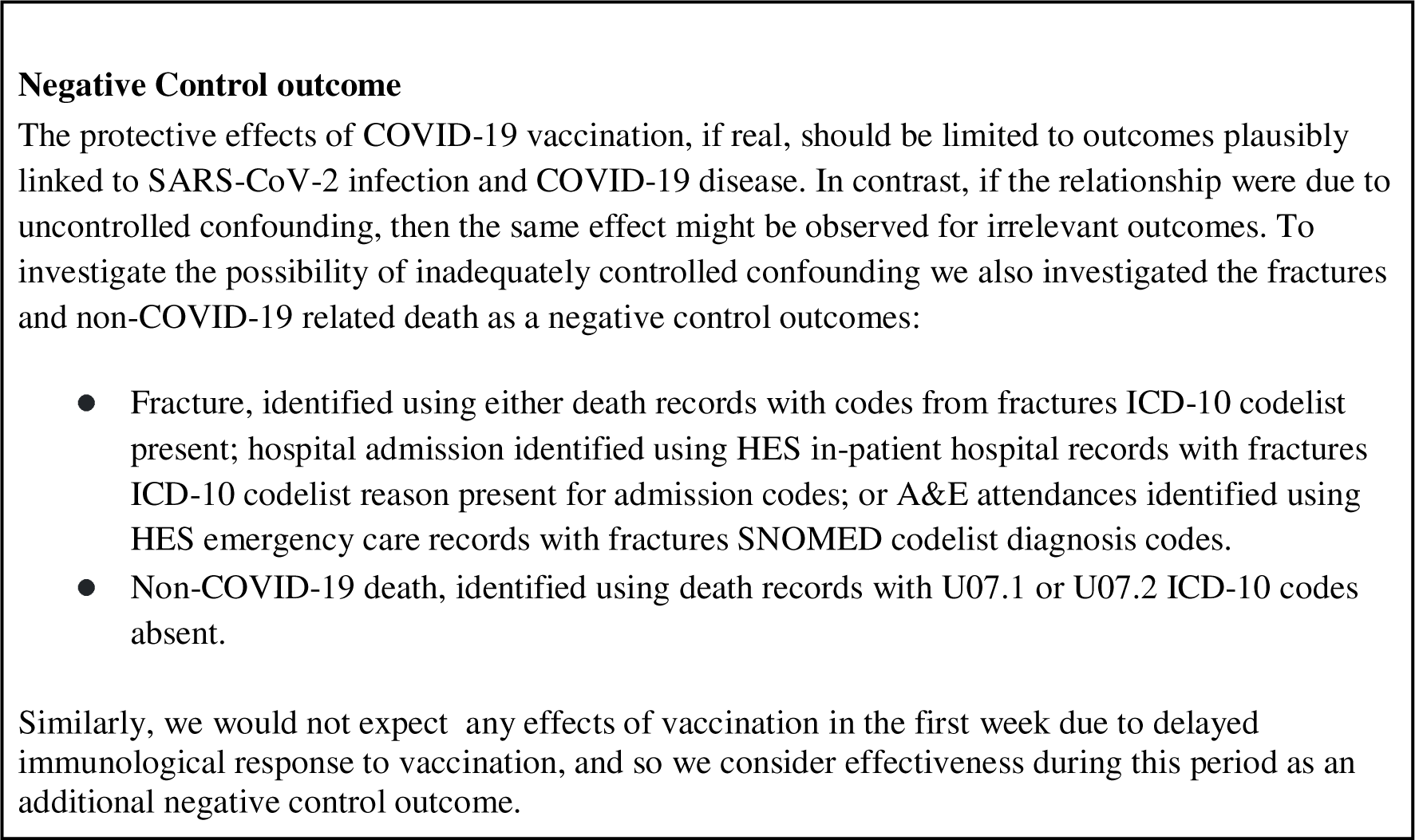
Definitions of Negative control outcomes.

