## Supplementary figures and images for "OpenSAFELY: Effectiveness of COVID-19 vaccination in children and adolescents"

### Supplementary Figure 1

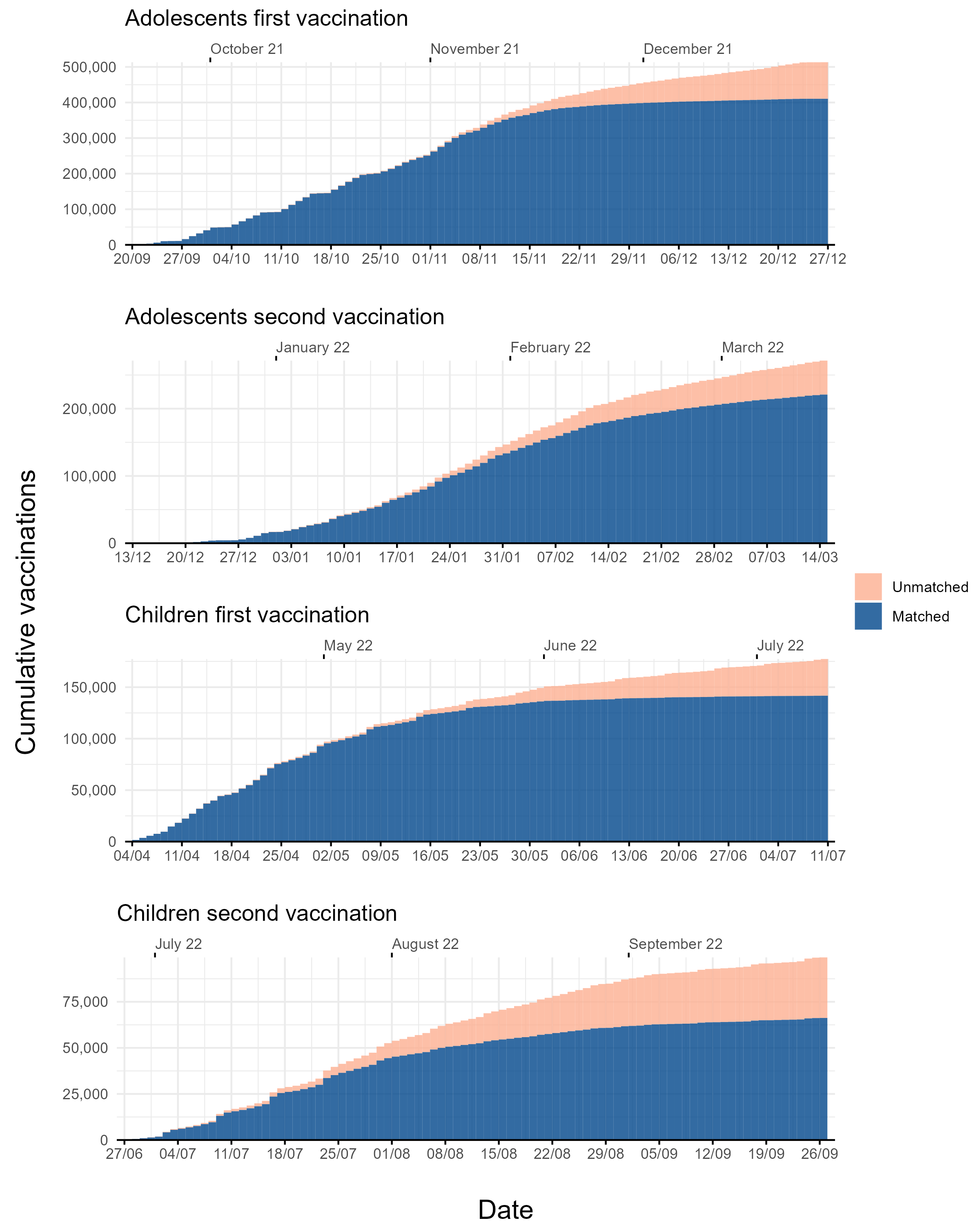

### Supplementary Figure 2

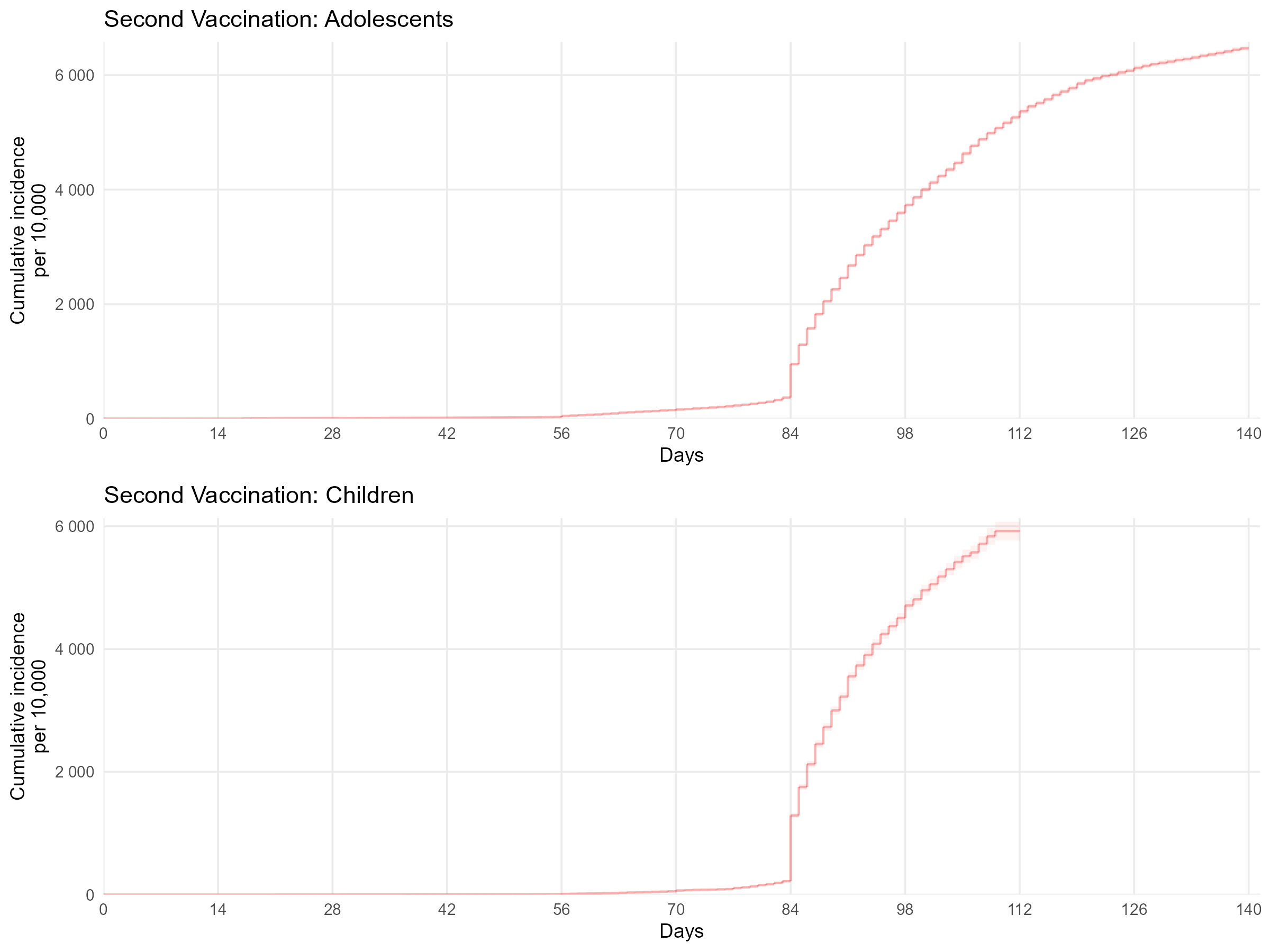

### Supplementary Figure 3

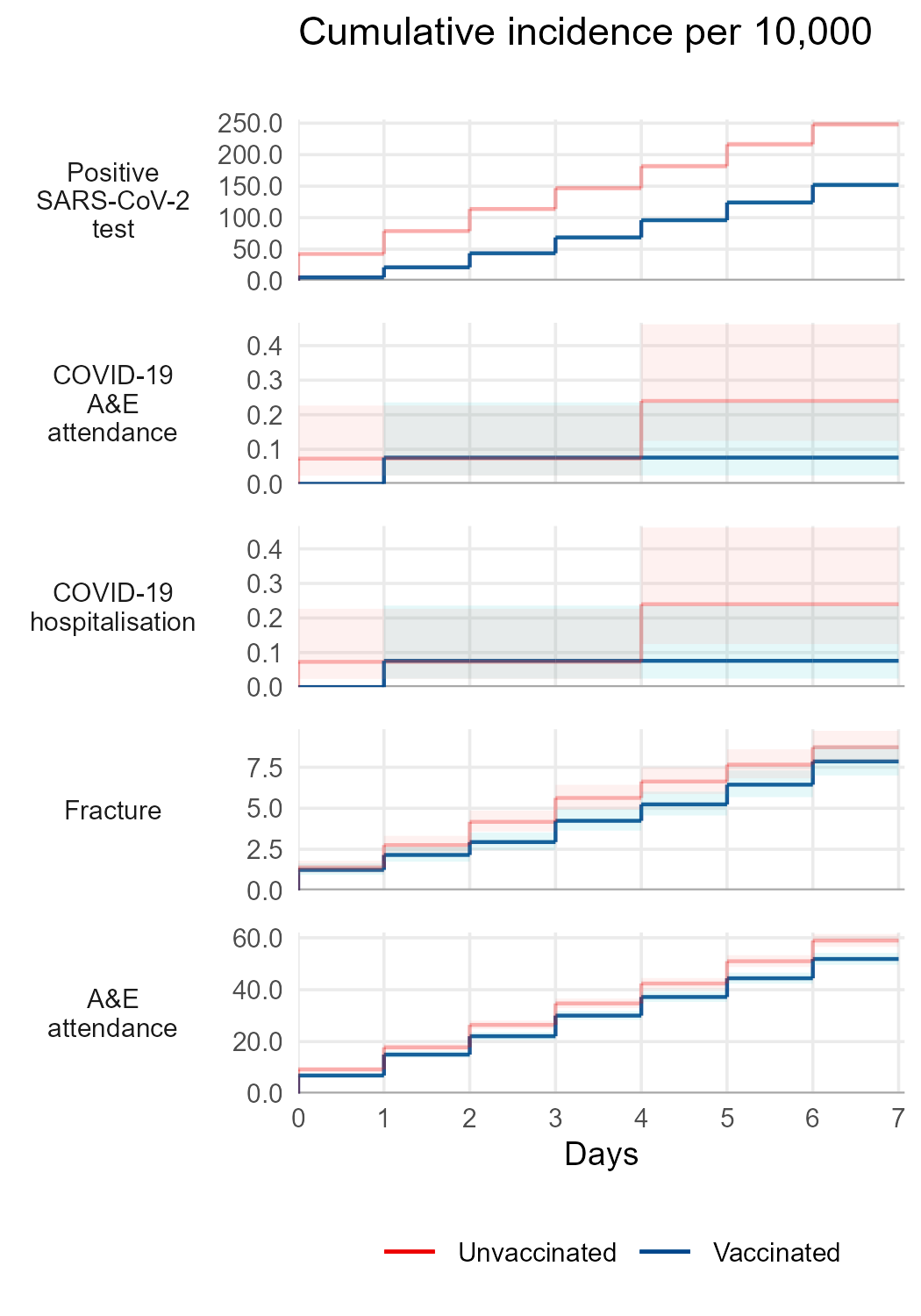

### Supplementary Figure 4

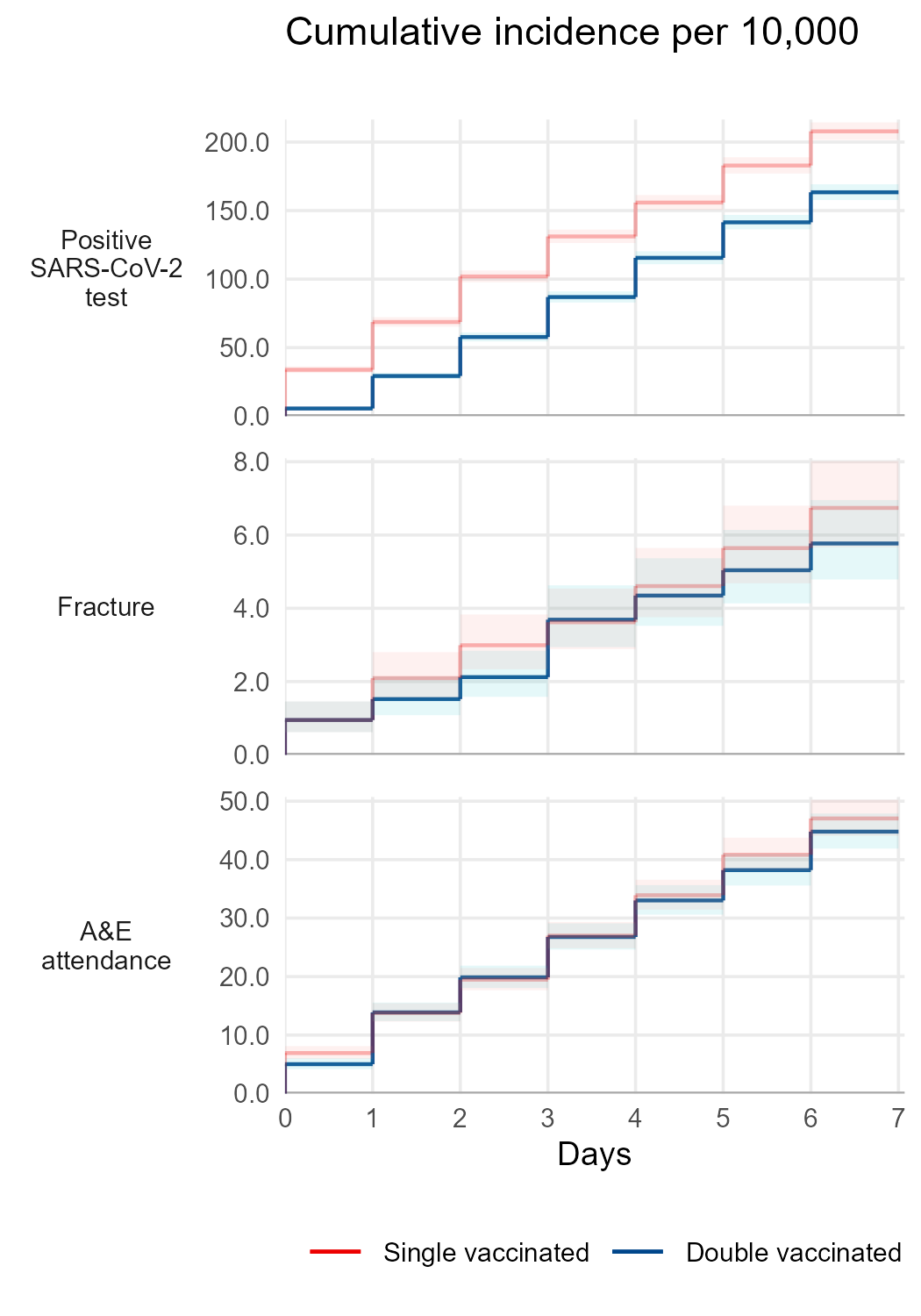
